# Effects of respite services on fluctuations in caregivers’ stress: A pilot study using a daily diary

**DOI:** 10.1101/2024.09.25.24314398

**Authors:** Koji Abe, Wataru Kubota

## Abstract

**Objectives:** Family caregivers for people with dementia report daily fluctuations in stress levels. Much prior research on caregivers’ daily stress fluctuations has used a daily diary method. However, only a few studies have used this method with family caregivers to examine the effects of respite services. This study aimed to use a daily diary method to assess the effects of respite services on stress and depression in family caregivers.

**Methods:** Participants included 13 family caregivers of persons with dementia using respite services in rural areas of Japan. Participants completed self-administered questionnaires every day for seven days, including the use or non-use of respite services, cognitive and daily living function, depression, and stress appraisal. Generalized linear mixed models with data nested within persons were used for the analysis.

**Results:** Few significant effects of services were found on caregivers’ depression and stress appraisals at the between-person level. However, within-person level analyses using generalized mixed models showed that respite services significantly reduced stress appraisal.

**Conclusion:** The findings demonstrate the stress-buffering effect of respite service for caregivers and the applicability of a daily diary method to the small sample.

## 1. Introduction

In recent years, the aging population has drawn increasing attention regarding the care of older adults. Caring for a person with a disability is stressful and challenging for families (Zarit et al., 1998). Stress accumulation in caregivers often leads to depression, so attempts to reduce stress and depression are being made around the world (Adams, 2008; Baumgarten et al., 2002). Providing respite services is one such program adopted in many countries to reduce stress and prevent depression in caregivers (Vandepitte et al., 2016). Although it is not possible to compare services between countries because the forms of respite and providers differ, respite services are defined as a supportive service provided in or outside the home to give the informal caregiver a temporary relief or break from caregiving duties (Vandepitte et al., 2016). Despite the potential advantages of respite care, research on its efficacy has been minimal, and a substantial body of empirical evidence on outcomes, either for caregivers or persons receiving respite, has yet to emerge (Zarit et al., 2017).

For example, Baumgarten et al. (2002) examined the impact of daycare services on the burden among family caregivers by randomized control trial but found no impact. Mossello et al. (2008) reported that providing respite services with informal counseling opportunities significantly reduced caregiver burden, although there was no impact on caregivers’ depressive symptoms. One reason for the discrepancy in these studies is that they relied on cross-sectional designs that emphasize between-person relationships or longitudinal designs that evaluate change over long periods (Mausbach et al., 2011). However, prior research has shown that the impact of daily stressors may be most significant on the day those events occur (Almeida, 2005). By interrupting the stress-overload cycle that is typically observed in people experiencing chronically high stress, respite services may make daily life more manageable, at least on days that caregivers receive respite (Zarit et al., 2014). Zarit et al. (2014), therefore, used a daily diary approach to examine fluctuations in stress and the effects of respite services. This approach enables the examination of between-person differences and within-person processes of change. The daily diary approach also indicated that a within-person perspective allows us to compare each person with themselves on respite and non-respite service days (Zarit et al., 2014; Zarit et al., 2017).

The current evidence suggests that respite services are an essential strategy for reducing caregiver stress, but more evidence is required on their effectiveness (Vandepitte et al., 2016). Only a few studies have used a daily diary with family caregivers. Therefore, as a pilot study, a small sample of participants was surveyed using a daily diary approach to examine the effects of respite services on daily fluctuations in caregivers’ stress and depression.

## 2. Methods

### 2.1. Participants

Questionnaires were distributed to 30 family caregivers of frail older adults who used respite services provided at a nursing home facility in October 2019. Thirteen consented to participate (consent rate: 43.3%), including two males and ten females (one did not respond). The mean age was 66.42 years (*SD* = 11.20). The care recipients included three males and nine females (one did not respond). The mean age is 86.17 years (*SD* = 7.67).

In this study, a questionnaire was administered for seven consecutive days. The facility staff gave participants a written explanation of the study, a consent form, and a questionnaire for seven consecutive days. Participants were then asked to complete the questionnaire every day after dinner but before bedtime. This study was conducted after obtaining approval from the ethical review committee of the Faculty of Law, Economics and Humanities, Kagoshima University (#10). Written informed consent was obtained from all subjects involved in the study.

### 2.2. Measurements

#### 2.2.1. Use of care services

To ascertain whether or not participants used respite services from Day 1 to Day 7, they were asked, “Today, did the older adults use respite services such as day services, daycare, or day rehabilitation?” The respondents answered “1. Yes” or “0. No.”

#### 2.2.2. Stress appraisal

We attempted to measure stress appraisals using a simple index to reduce the burden on respondents. The item indicating overall burden, “Overall, how stressed do you feel in caring for your relative?” was developed as a one-item stress appraisal of caregiving. The item was scored on a five-point scale from “1. not at all stressful” to “5. very stressful,” with a higher score indicating more significant stress.

#### 2.2.3. Depression

The Japanese version of the K-6 (Furukawa et al., 2008) was used to measure depression. Participants rated depressive symptoms using a five-point scale. Higher scores indicate more depression. The lowest internal consistency was α = 0.78 on Day 1, and the highest was α = 0.98 on Day 5.

#### 2.2.4. Cognitive and daily living functions

The Dementia Assessment Sheet for Community-based Integrated Care System 8-items: DASC-8 by Toyoshima et al. (2018) was used to assess the cognitive and daily living functions of older adults needing daily care. Higher scores indicate a decline in cognitive function, Activities of Daily Living (ADL), and Instrumental Activities of Daily Living (IADL). In this study, the scale scores used for analysis were calculated by summing the scores of all items. Internal consistency was high in this study (α = 0.88). The average score for the DASC-8 was 23.63 (*SD* = 5.95), and 11 out of 13 family caregivers cared for older adults with moderate or severe disabilities.

### 2.3. Analysis

Generalized Linear Mixed Models (GLMM) were used to analyze daily stress fluctuations of individual levels in caregivers. In the GLMM, stress and depression were set as dependent variables, and the intercept was whether or not respite services were used (binary). The cognitive and physical functions of older adults that were measured on the first day of the survey, the gender of the caregiver (binary), and the age of the caregiver were set as fixed effects. The variance and covariance matrix for the random effects was unstructured, assuming no specific structure.

In a previous study, Zarit et al. (2014) measured daily fluctuations in Behavioral and Psychological Symptoms of Dementia (BPSD) and examined the effects of these fluctuations on mental and physical health. However, we anticipated a more significant burden on responses about BPSD each day and the possibility of increased missing data. Therefore, we only measured older adults’ cognitive and physical functioning on the first day. The DASC-8 is considered a relatively stable factor, such as daily care needs and memory function decline, rather than an unstable factor, such as the BPSD. IBM SPSS Statistics version 27.0.1 for Windows was used for all data processing and statistical analyses.

## 3. Results

### 3.1. Descriptive statistics

Figure 1 shows the daily mean change in stress appraisal and depression. The highest stress appraisal was on the first day, with a mean of 2.62 (*SD* = 1.19). The lowest stress appraisal was on the fifth day, with a mean of 1.73 (*SD* = 1.27). For depression, the highest mean was on Day 3 with a mean of 9.00 (*SD* = 6.99), and the lowest was on Day 6 with a mean of 4.90 (*SD* = 5.23). There was no significant correlation between the DASC-8 score on the first day and stress appraisal or depression.

**Figure 1.**
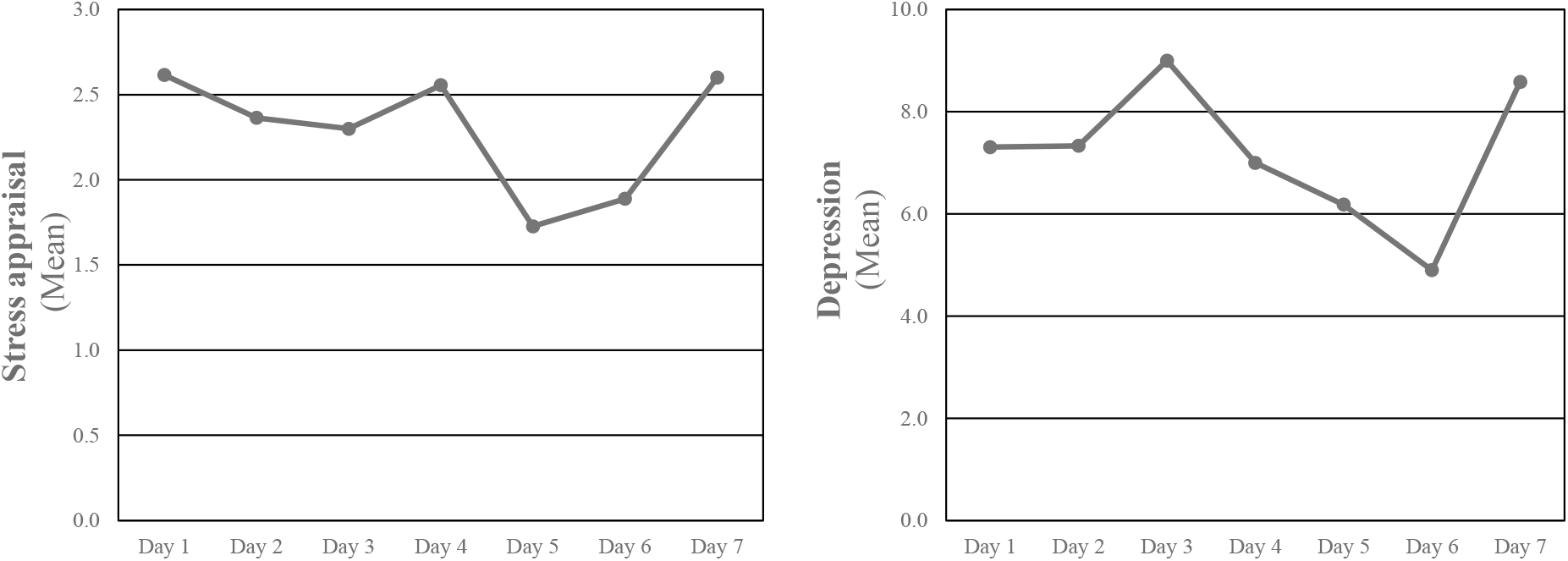
Daily fluctuations in stress appraisal and depression among family caregivers

### 3.2. Daily effects of service use on stress and depression

Next, we compared stress appraisal and depression with the use of respite services on each day. There were no significant between-person differences for almost all days except on Day 7, where there was a significant difference in stress appraisal (*F* (1, 8) = 10.82, *p* = .011)

### 3.3. Effects of service use on daily fluctuations in stress and depression

The GLMM (Table 1) showed significant within-person effects of service use on caregivers’ stress appraisal (coefficient estimates = -0.774, *p* = .006). The intercept was also significant. On the other hand, no significant associations were found between stress appraisal and caregivers’ gender, age, and older adults’ cognitive and physical functions.

**Table 1.** Generalised linear mixed models for daily stress appraisal and depression.

|  |  | Stress appraisal |  |  | Depression |  |  |
| --- | --- | --- | --- | --- | --- | --- | --- |
|  |  | Coefficient | SE | p-value | Coefficient | SE | p-value |
| Fixed Effects |  |  |  |  |  |  |  |
| Intercept |  | 6.939 | 3.198 | 0.034 | -2.404 | 24.618 | 0.923 |
| Service Use | Yes | -0.774 | 0.272 | 0.006 | -1.573 | 1.178 | 0.186 |
|  | No | Reference |  |  | Reference |  |  |
| Caregiver's Gender | Female | -2.031 | 1.111 | 0.072 | 1.364 | 8.503 | 0.873 |
|  | Male | Reference |  |  | Reference |  |  |
| Caregiver's Age |  | -0.067 | 0.037 | 0.075 | 0.033 | 0.282 | 0.906 |
| Cognitive and daily living functions |  |  |  |  |  |  |  |
| DASC-8 |  | 0.086 | 0.053 | 0.111 | 0.318 | 0.383 | 0.409 |
| Random Effects |  |  |  |  |  |  |  |
| Intercept VAR |  | 0.643 | 0.451 | 0.154 | 46.756 | 25.408 | 0.066 |
| AIC |  | 220.493 |  |  | 435.039 |  |  |

Notes: DASC-8: dementia assessment sheet for community-based integrated care system 8-items, VAR: variance, AIC: Akaike information criterion, SE: standard error

In the analysis with depression as the dependent variable, no significant effects were found. When each model was evaluated by the Akaike Information Criterion (AIC), the analysis with stress appraisal as the dependent variable was lower than that with depression.

## 4. Discussion

Results of descriptive statistics show considerable fluctuations in daily stress appraisal and depression. Our results indicate high stress or depression on a given day, and it may not also occur on the next day. Therefore, the timing of measurement may significantly influence a diagnosis.

As indicated in this study, stress fluctuates daily, and it is likely that during a given week, there may be days in which caregiving for older adults is difficult and other days in which it is not. In order to identify the factors related to caregivers’ stress, it is important to conduct an intensive longitudinal study rather than a cross-sectional survey (Gérain et al., 2023).

The results of this study may also indicate the difficulty in dealing with the impact of service use on stress appraisal and depression. When we examined the use of services from Day 1 to Day 7, no other significant between-person differences were found, except for stress appraisal on Day 7. Although many reports have examined the effects of using respite services on caregivers and older adults, their results are not easy to interpret because older adults’ disabilities, the period of caregiving, and communication with service staff may differ from person to person. Our cross-sectional analysis showed little between-person differences of stress appraisal and depression due to service use, suggesting that individual differences in service effectiveness are highly relevant.

The GLMM in this study showed that respite services affected stress appraisal at the within-person level but not depression. Zarit et al. (2014) found the direct effects of services on stressors and indirect effects on depressed or positive caregiver experiences. Gaugler et al. (2003) also reported using respite services reduced feelings of role overload of caregivers and memory problems of older adults. The results of the present study, like those of the earlier report, seem to imply that respite services do not directly improve caregivers’ depression but indirectly improve it through a buffering effect for stress appraisal.

### 4.1. Limitations

This study was a pilot study, and many issues remain to be addressed. First, the most significant issue is the sample size. Although 30 people were asked to participate in this study, only 13 volunteered due to the burden of responding to the survey every day for a week. Due to the small sample size and low participation rate, it is difficult to say that similar results would be obtained in other samples. However, several previous studies using intensive longitudinal methods have reported results with fewer than 20 participants (Fonareva et al., 2012; Jain et al., 2014; Konnert et al., 2017; Rullier et al., 2014). Along with these reports, this study demonstrated that a daily diary method is effective with caregivers. In the future, it will be necessary to conduct a more comprehensive survey that includes caregivers’ stress, depression, and the fluctuation of BPSD to capture the daily fluctuation of caregivers’ stress and mood.

Second, this study examined the effect of respite services only on whether or not they were used; the contents of these services are unknown. In Japan, respite services are provided through a combination of daycare, day rehabilitation, home help, and home rehabilitation, making it difficult to identify which specific service affects caregiving stress.

Third, caregiver respite is not provided only through formal services. For example, assistance from family members who do not reside with caregivers, volunteers, and community members has been identified as a factor that can reduce stress. It is considered necessary to examine the details of respite services and informal support to develop a supportive community for caregivers.

## 5. Conclusions

This study examined the effects of respite services on daily fluctuations in stress and depression among family caregivers. Although the sample size was small, the results indicated that respite services significantly reduced caregivers’ stress appraisal. These results indicated the stress-buffering effect of respite service for caregivers and the applicability of a daily diary method to the small sample.

## Data Availability

All data produced in the present study are available upon reasonable request to the authors.

## Author Contributions

Koji Abe: Conceptualization, Methodology, Software, Validation, Formal analysis, Investigation, Resources, Writing-Original Draft, Writing-Review & Editing, Project administration, Funding acquisition.

Wataru Kubota: Conceptualization, Investigation.

## Data Availability Statement

The data will not be shared because of confidentiality based restrictions imposed by ethical review committee of the Faculty of Law, Economics and Humanities, Kagoshima University, unless requested through an administrative procedure.

